# Fine-Grid Spatial Interaction Matrices for Surveillance Models, with Application to Influenza in Germany

**DOI:** 10.1101/2025.04.03.25325159

**Authors:** Manuel Stapper, Sebastian Funk

## Abstract

Accurate spatial forecasting of infectious disease outbreaks is critical for public health planning, yet models often rely on administrative boundaries that may not reflect true transmission patterns. We propose the Fine-Grid Spatial Interaction Matrix (FGSIM), which models transmission risk between districts based on distances between individuals to improve both forecast accuracy and spatial resolution of risk mapping. FGSIM creates district-level transmission matrices by defining individual-level transmission risk and aggregating to the district level. We transformed five distance metrics into contact intensity measures to create weight matrices within the endemic-epidemic modelling framework. We evaluated FGSIM using German influenza data (2001-2020) and compared its performance to established methods across four study regions for one to eight weeks ahead forecasts. FGSIM outperformed simplified variants and models without spatial dependence in most cases when evaluated in-sample. For one-week-ahead forecasts, a centroid-based model performed best in three of four regions. For longer-term forecasts, a circle-population based model consistently outperformed others. Risk maps at 100m resolution demonstrated FGSIM’s ability to identify high-risk areas not aligned with administrative boundaries. FGSIM provides a flexible, computationally feasible approach to incorporating individual-level risk into district-level infectious disease models, showing competitive performance in forecasting and enabling fine-scale risk mapping for targeted public health interventions.

**Availability:** All data used are publicly available and summarised at https://zenodo.org/records/15600537. General functions to apply the methods are available in a toolbox at https://github.com/ManuelStapper/ FGSIM. Application-specific code is available at https://github.com/ManuelStapper/FGSIM_Application. **Contact:**

**Supplementary information:** Supplementary data are available at *Journal Name* online.

## 1. Introduction

Infectious diseases are transmitted through complex networks of interactions between individuals. While many factors influence contact patterns, such as social networks, workplace environments, and surrounding infrastructure, spatial proximity between an infected and a susceptible individual is especially important. The underlying assumption is that diseases tend to spread more easily between geographically close regions or individuals due to the higher frequency of interactions that can lead to transmission.

One of the aims of spatial modelling in epidemiology is to quantify and understand the intensity of contact and thus transmission between geographical units. Capturing these dependencies allows for more accurate predictions of disease spread and provides insights into its underlying dynamics. However, modelling spatial dependence often requires a trade-off between simplicity and complexity. Simple models make strong assumptions about human behaviour, which makes them easy to implement and maintain and computationally efficient but limits their ability to capture critical factors influencing disease transmission. In contrast, complex models can incorporate detailed human movement patterns but demand extensive data and computational resources. An important challenge lies in making realistic assumptions about human movement while ensuring computational feasibility of fitting the model.

A straightforward measure of spatial proximity is to define the closeness of relevant areas (e.g. administrative districts) as the distances between their centres. From this starting point, the intensity of interaction between districts can be modeled as a function of those distances. For instance, one could assume a constant interaction intensity for the nearest districts or use a decay function to model how contact intensity decreases with distance. Another commonly used approach is defining closeness based on the order of neighborhood, i.e., the minimum number of district boundaries an individual must cross to move between the districts. Contact intensity can then be specified using this measure; for example by considering only first-order neighbors or by assigning equal interaction intensities to districts within the same order of neighborhood. A widely used method, rooted in the work of Brockmann et al. [2006], applies a power law to the order of neighborhood, assuming that contact intensity decays geometrically with increasing distance [Meyer and Held, 2014]. While practical and effective, these models depend on area boundaries and are sensitive to where they are. See Appendix A for a discussion on the sensitivity of the order of neighbourhood.

For modelling the probability of contact between two individuals, the distance, whether expressed as Euclidean distance or neighborhood order is not the only determining factor. Individuals living in densely populated districts are less likely to travel long distances than those in remote areas. Similar to Newton’s law of gravitation, Zipf [1946] proposed that human movement follows a ”gravity model”, where the probability of an individual traveling to a destination depends on the populations at the origin and destination and decreases with increasing distance between them [see, for example, Haynes and Fotheringham, 1985].

Other approaches incorporate factors beyond distance to model human movement. Stouffer’s law of intervening opportunities states that migration is proportional to the opportunities at the destination and inversely proportional to those between origin and destination [Stouffer, 1940]. This principle motivates the radiation model [see, for example, Simini et al., 2012, Alis et al., 2021], which considers populations at the origin and destination while accounting for the population in the circle around the origin that touches the destination. For a detailed overview of human mobility models, see Barbosa et al. [2018] and Tizzoni et al. [2014].

Most of the models discussed above operate at the level of administrative geographical units, aggregating population movement and interaction patterns. However, this level of generalization can be relaxed to increase model flexibility. Agent-based models, for instance, define movement and interactions at the individual level rather than the district level.

The remainder of this article introduces a novel approach to spatial dependence modelling that defines interactions at the individual level and then aggregates them to the level of administrative geographical units (henceforth, “districts”). In this framework, individual interactions are treated as random variables, where stochasticity arises from the randomly selecting individuals from districts. Aggregation to the district level is performed by taking expectations. This method combines the advantages of individual-level modelling with the simplicity of district-level modelling. By focusing on individual locations rather than predefined district boundaries, this approach avoids restrictive assumptions about human movement patterns, while maintaining computational feasibility.

## 2. Methods

### 2.1. Model

We applied the endemic-epidemic framework introduced by Held et al. [2005] to model the number of newly infected individuals in spatial units. Its implementation in the *surveillance* R package is widely used for spatio-temporal epidemic data. This versatile framework partitions the expected number of new infections into endemic and epidemic components [Meyer et al., 2017].

Let *Y_it_* be the number of new cases in district *i* at time *t* and *Y_t_* the vector of observations of all areas *i* = 1*, …, N*. Given the information set available at time *t* − 1, i.e. the sigma algebra F*_t−_*_1_ generated by {*Y_t−_*_1_*, Y_t−_*_2_*, …*}, the expectation *µ_it_* = E(*Y_it_*|F*_t−_*_1_) is defined as

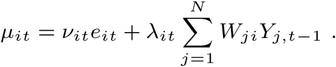

The first term captures the baseline infection counts, explained by usual circulation. It depends on *e_it_*, the number of (susceptible) individuals in district *i* at time *t*. The second term, the epidemic component, captures the transmission of cases, with W denoting the weight matrix. Each element *W_ji_* represents the intensity of contacts between units *j* and *i* that potentially lead to a transmission of the disease from *j* to *i*. The weight matrix is typically row-normalised allowing each row *W_j·_* to be interpreted as the distribution of contacts from an infected individual in *j* across all districts.

Both components are scaled by log-linear predictors *ν_it_* and *λ_it_*, defined as

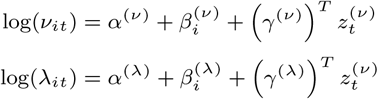

Here, *α*^(*ν*)^ and *α*^(*λ*)^ are intercept parameters, *β_i_* are district-specific effects, which can be either random or fixed effects. Other covariates *z_t_*may include seasonality components and *γ^ν^* and *γ*^(*λ*)^ are the corresponding parameter vectors. To ensure identification, the intercept parameters *α*^(*ν*)^ and *α*^(*λ*)^ are often dropped in presence of district effects or the average district effect is set to zero.

The number of new infections is then assumed to follow a Negative Binomial distribution:

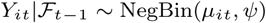

where the Negative Binomial distribution can be replaced by a Poisson distribution or a Negative Binomial with district-specific overdispersion parameters *ψ_i_*. For additional flexibility in modelling intra-district transmission, the epidemic term is often decomposed into an intra-district (autoregressive) component and an inter-district (neighbour-driven) component.

In the remainder of this article, we focus on the weights *W_ji_*, which capture the dynamics of transmission. A well-established approach is to define weights using the neighbourhood order between districts *O_ji_* and applying the power law such that unnormalised weights are given by 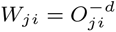, with *d* as a decay parameter.

Our proposed alternative for defining weights is based on a similar principle, but uses a distance measure at the individual level. For instance, consider a pair of districts *i* and *j*. We select an individual from each district at random and measure the distance *D_ji_* between them (e.g. the beeline distance). Assuming that the probability of contact decays with distance according to a power law, the contact intensity is proportional to 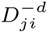. However, since *D_ji_* is a random variable due to the random selection of individuals, the individual transmission risk is a random variable as well. Hence, we aggregate the individual transmission risk by taking the expectation 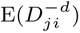, where the expectation is taken with respect to the distribution of individuals living in the two districts. Computing 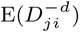 exactly is computationally infeasible when considering all pairs of individuals. Instead, assumptions are made about the distribution of *D_ji_*.

This process involves four steps. First, locations of individuals from each district are repeatedly sampled according to the population density. Second, distances between the sampled pairs are calculated, yielding a sample of distances. Third, these distances are used to fit a parametric distribution. Finally, the fitted distribution is used to translate distances into weights by taking expectations.

The Lognormal distribution is a natural distributional assumption for modelling *D_ji_*, as it takes only positive values and has desirable properties. Specifically, if *D_ji_*is Lognormal, 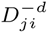 also follows a Lognormal distribution, allowing for a closed-form expectation. However, the Lognormal distribution may not adequately account for bounded distances due to district boundaries and may be overly restrictive. To address this, we assume that *D_ji_* follows a mixture of truncated Lognormal distributions. This approach preserves the closed-form expectation 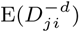 while providing greater flexibility and ensuring boundedness. The mixture is fitted using an Expectation-Maximisation (EM) algorithm, where we used the Bayesian Information Criterion (BIC) for the selection of the number of mixing components in order to avoid overfitting [Schwarz, 1978].

In practical applications, truncating the power law can be advantageous to prevent extreme values for small distances. Further, it is a reasonable assumption that individuals have a small radius around their home with uniform movement. To account for this, we generalise the power law by introducing a truncation threshold, *δ >* 0 such that the weights are defined as the minimum of 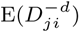 and *δ^−d^*. The truncation parameter *δ* is estimated alongside the other model parameters. For a full derivation of the closed-form expectations, see Appendix Balong with derivatives of these expectations, required for numerical optimisation.

We applied five distance measures in the new approach, in each case first sampling a large number of individuals per district according to population density. Data on population distributions within districts is available from WorldPop [2018] on a 100m by 100m grid.

#### Beeline Distance

The simplest and most intuitive measure of distance between individuals is the geodesic, or beeline, distance. This measure is easy to compute and provides a baseline for comparison with more complex alternatives. It was calculated using the Haversine formula, as implemented in the *geosphere* R package [Hijmans et al., 2023].

#### Travel Time

While the beeline distance serves as a straightforward starting point, it may not reflect real-world travel patterns. For example, two locations separated by a river may have a short beeline distance but require a significantly longer journey via a bridge. Additionally, regions with high volumes of traffic often have well-developed infrastructure, such as highways, which influence travel behaviour. These factors make travel time a more realistic measure of the likelihood of interaction between individuals.

Travel times were calculated using the *osrm* R package [Giraud, 2022], which computes road travel times based on OpenStreetMap data. This package supports various modes of travel; we used car travel times. Although travel time provides valuable additional information, it is computationally more demanding than the beeline distance due to the need for route calculations.

#### Gravity Model

Individuals are more likely to travel to areas with higher population densities, reflecting the gravitational pull of urban centres. The gravity model incorporates this principle by combining the beeline distance between two locations, *D_ji_*, with population densities at the origin (*P_j_*) and destination (*P_i_*):

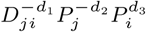

Assuming the triple (*D_ji_, P_j_, P_i_*)*^′^* is jointly Lognormal, this formula has a closed-form expectation. However, assuming (mixtures of) truncated Lognormal distributions for at least one element of the triple jeopardises the closed-form expectation. Further, preliminary analyses suggest that *P_i_*and *P_j_* are not well-approximated by a mixture of bivariate Lognormals. To simplify the model, we assumed equality of *d*_2_ and *d*_3_. Hence, we included the population ratio *P_ji_* = *P_j_/P_i_* which fits the Lognormal assumption more closely. Instead of the triple above, we measured the distance as (*D_ji_, R_ji_*)*^′^* and translated it to a contact intensity 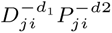.

#### Circle Distance

The circle distance expands on the gravity model by considering the population within a circular area around the origin. For a given destination, the measure accounts for all inhabitants within a circle centred at the origin and touching the destination. This approach captures the attractiveness of a destination relative to alternative locations within the same range.

Using high-resolution population data on a 100m grid enables precise calculation of circle populations, but the process is computationally intensive. For efficiency, we used different resolutions depending on the proximity of locations: a 100m grid for intra-district distances and a coarser 1km grid for inter-district distances. We adjusted the population counts to account for discrete approximation of the circles using grid cells.

#### Radiation Model

The radiation model builds on the circle distance by incorporating the populations at both the origin and destination. It evaluates transmission risk using the following formula:

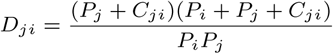

where *C_ji_* is the population within a circle around the origin that touches the destination. Traditionally used to estimate flux between locations in non-parametric settings, the radiation model here serves as a distance measure. It balances simplicity with the ability to account for population distributions, making it a natural competitor to the gravity model.

#### Simplifications

In addition to above weight matrix formulation, we added two levels of simplification. First, we added a version where transformation and expectation were inverted. For the beeline distance measure *D_ji_*, the weights are E(*D_ji_*)^−*d*^ instead of 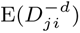. The resulting weights lose their interpretation as an average transmission risk between individuals. Instead, they can be seen as a transmission risk between average individuals of districts. The advantages are lower computational complexity as well as less memory needed to store the expectations E(*D_ji_*) compared to storing parameters of a mixture distribution. Although this simplification is feasible for all distance measures, we only included it for the beeline distance and the gravity distance. For the latter, the reformulation is similar and transforms expected distance and the expected population ratio to a transmission risk.

This simplification still uses detailed population density data. An additional step of simplification is ignoring population density data and base weights on the centroids of districts. We applied the power law to the distance between centroids to quantify transmission risk. We included weights based on the beeline distance between centroids as well as a centroid gravity model that accounts for the total population in districts. Similar to above fine grid approach, we defined those centroid gravity weights as product of power laws applied to the beeline distance and the population ratio. To maintain the ability to express intra-district transmission risk, we introduced a new parameter for this approach such that the raw weights are *W_ji_* = *ω >* 0 for *j* = *i*. The approach is equivalent to extending the endemic epidemic model to three components where the autoregressive and neighbourhood epidemic component have equal loglinear predictors.

### 2.2. Application

We applied the new approach to influenza cases in Germany from 2001 to the 45th week of 2020. The data, sourced from Robert Koch Institute [2024], contained the number of weekly confirmed infections with influenza virus reported for the 401 administrative districts. The observation period overlaps with two relevant epidemic outbreaks: the 2009 swine-flu outbreak and COVID-19.

We excluded data from the period of the H1N1 pandemic, the 52 weeks starting from the 17th week of 2009, from the analysis as these cases likely did not follow with the transmission mechanisms captured by the model. The starting point of this exclusion period was chosen based on the lowest infection count between seasonal peaks. We ignored the overlap with COVID-19 pandemic as the first minor outbreaks in Germany started at the beginning of 2020 when influenza infections had already declined to endemic levels.

The 401 districts were grouped into four regions (north, east south and west), which served two purposes:

*•* Reducing computational complexity, as the number of transmission paths grows quadratically with the number of districts.
*•* Exploring how model performance varies across regions with distinct spatial and demographic characteristics.

The northern region included Schleswig-Holstein, Hamburg, Bremen, and Lower Saxony. The east was defined as Berlin, Brandenburg, Mecklenburg-Western Pomerania, Saxony, Saxony-Anhalt and Thuringia. The south comprised Bavaria and Baden-Württemberg, and all remaining states form the west. Figure 1 provides an overview. These regions differ significantly in characteristics such as population size, urbanisation, and spatial distribution. For example, the northern region includes sparsely populated rural areas as well as isolated urban centres like Hamburg and Bremen. The west exhibits a cluster of densely populated areas in the Ruhr region. The largest districts by area are sparsely populated and located in eastern Germany alongside the largest district by population: Berlin. In the south, we observed many nested districts, where urban districts are surrounded by their corresponding rural district.

**Fig. 1:**
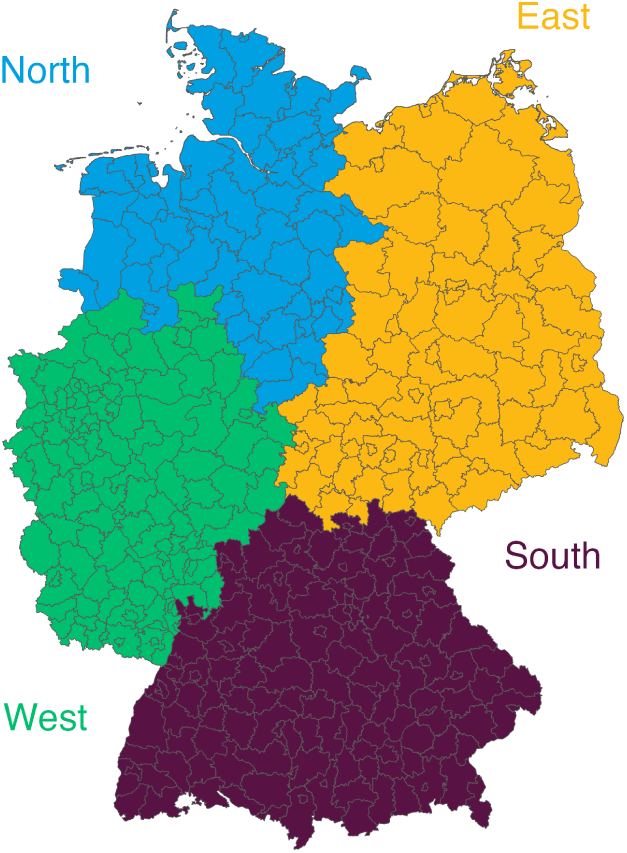
Spatial division of Germany’s 401 administrative districts into four regions

For each of the four regions, we compared eleven ways to define spatial dependence. As a baseline, the first model assumed no spatial dependence, i.e. a diagonal weight matrix. The second is the power law approach applied to the order of neighbourhood. These approaches will be abbreviated as None and NGB in the following. Further, we included the five distance measures of the fine grid approach introduced in the previous section based on the beeline distance (FG-B), the travel time (FG-T), a gravity attraction (FG-G), the circle population (FG-C) and the radiation formula (FG-R) respectively. Simplified versions of the beeline and gravity approach are denoted as FGs-B and FGs-G for the simplified fine grid approach and Cent-B and Cent-G for the centroid based weights.

We evaluated these approaches in terms of their in-sample performance as well as in a forecast study. We assessed out-of-sample performance for a rolling window of around 18 years of data for estimation and forecasting up to 8 weeks into the future before shifting the window by one week. This resulted in around 100 time windows, or two years.

To be able to use the FG approach, we needed to fit a mixture of (truncated) LogNormal distributions for each distance measure and each pair of districts that was in the same region. We did this by carrying out the following steps:

*•* Sample 2000 pairs of individuals, one from each district, weighted by the population density.
*•* Compute the distance measure for each pair.
*•* Fit a mixture of truncated Lognormal distributions to the sample of 2000 distances using the EM algorithm, where the number of mixture components is determined by information criteria and its upper bound set to five components.
*•* Saving the estimated parameters and mixing weights.

Sampling 2000 individuals per districts resulted in 802,000 individuals in total, or around 1% of the total population. We found the number of samples sufficient to obtain stable estimates of LogNormal parameter estimates while maintaining computational feasibility. Figure 5 in the appendix summarises the number of mixing components. We saw that the upper bound of five components sufficed. The radiation model tended to have few components. For all four regions, the distribution of radiation distances was well approximated using only one component for most pairs of districts with an average of 1.5 components. The bivariate gravity distance measure needed most mixing components with 2.7 on average.

We used similar model architecture across all four regions and for each distance measure. We included fixed effects for the districts in both the endemic and epidemic component. In the endemic component, we further assumed constant population sizes, such that the component reduced to *ν_it_*. We included harmonic waves with one year periodicity in order to capture seasonality. The number of harmonic waves for each distance measure was determined in a forward selection according to the BIC in the application to the training data that was not predicted in the forecast performance assessment. Starting with a model that omitted seasonality terms, we increased the order of the harmonic waves by one, separately in the endemic and epidemic component, and determined the increase to be selected by the largest decrease in BIC. We stopped the stepwise selection if an increase of the seasonality orders did not improve the BIC

We assumed overdispersion parameters to be the same for all districts in a region.

For each distance measure, we explored four variations: with and without population scaling, i.e. accounting for heterogeneity in population sizes of districts at risk, as well as with or without truncating the power law for small distances.

After fitting a model, we generated a total of 1000 trajectories of length 8 weeks into the future following the fitted model. From this approximation of the predictive distributions, we saved quantiles (2.5%, 5%, 10%, 50%, 90%, 95% and 97.5%) and scored the prediction using the weighted interval score (WIS) and the ranked probability score (RPS). We computed relative scores as the ratio of an individual model’s score and the score of the reference model with no spatial autocorrelation.

We aimed to assess the usefulness of the new method in three areas: additional insight into the dynamics of disease transmission, forecast performance, and a more detailed spatial risk assessment.

### 2.3. Risk Mapping

The fine-grid sptaial modelling approaches incolved defining distances at the individual level before aggregating transmission risks to the district level. Once a model is fitted, this process can be reversed, allowing us to derive the expected number of new infections at a finer resolution than districts to give a detailed risk map.

We determined the expected number of new infections in a grid cell by separately calculating the expected number of new endemic and epidemic infections. For endemic infections in a district *i*, the expected number was given by *ν_it_*, which was then distributed among the district’s grid cells in proportion to their population density. We calculated the expected weekly epidemic infections by computing the expected number of secondary infections in every grid cell caused by one infected individual residing in that district. To do so, we iterated over all grid cells within the district of origin and calculated distances to all other grid cells within the study area. We translated these distances into contact intensities by applying the power law. We then compute a weighted average where the weights were proportional to the population in the grid cell of origin. This yielded the contact intensity from the district of origin to all grid cells. We normalised the contact intensities to represent the probability distribution of expected contacts from one person in the district of origin to all other grid cells. Scale these probabilities by the observed number of cases in the district during the current week and sum them across all districts. The resulting value represents the aggregated risk from all observed cases across districts, which is then scaled by the corresponding epidemic log-linear predictor *λ_it_*.

By summing the expected endemic and epidemic infections, we obtained the total risk for each grid cell. This risk measure highlights areas with a high expected number of cases in absolute terms, correlating with population density. To derive an individual risk map, which reflects the per-person risk of infection, we divided the total risk in a grid cell by the number of residents.

## 3. Results

### 3.1. In-Sample Performance

The novel method provides flexibility in defining weight matrices. To narrow down the selection of models included in the rolling window forecast evaluation, we began by inspecting in-sample performance. In a preliminary analysis, we observed that scaling raw weights by population sizes resulted in a lower BIC for all distance measures and across all four regions. On the other hand, truncating the power law — and thus introducing an additional parameter — did not consistently reduce the BIC. In cases where truncating the power law did reduce the BIC, the improvement was small. Therefore, for the following analysis, we focus on models with population scaling and an untruncated power law.

Table 1 summarises the selected seasonality orders and the BIC from fitting to the complete data set. The seasonality orders differed slightly between regions and distance measures, but were two or three except for Northern Germagrouny. According to the BIC, the centroid gravity model had the best in-sample fit for Northern Germany. In the other three regions, the radiation approach outperformed the other approaches. In terms of levels of simplicity, we saw that no simplification dominated the two other approaches when using the beeline distance. With a gravity distance measure, both simplifications had a lower BIC compared to the Fine Grid approach. In all other regions, the Fine Grid approach had a lower BIC than the simpler versions.

**Table 1.** Selected seasonality order (Endemic, Epidemic) and the corresponding BIC from fitting to complete data.

| Model | North |  | East |  | South |  | West |  |
| --- | --- | --- | --- | --- | --- | --- | --- | --- |
|  | Order | BIC | Order | BIC | Order | BIC | Order | BIC |
| None | (2,2) | 85,972 | (3,3) | 143,807 | (2,3) | 220,688 | (3,3) | 162,591 |
| NGB | (1,3) | 80,144 | (3,2) | 135,720 | (3,3) | 204,397 | (3,2) | 150,741 |
| FG-B | (1,2) | 79,965 | (3,2) | 135,539 | (3,3) | 204,174 | (3,2) | 150,306 |
| FGs-B | (1,3) | 80,197 | (3,2) | 136,008 | (3,3) | 204,833 | (3,2) | 150,701 |
| Cent-B | (1,2) | 80,003 | (3,2) | 135,915 | (3,3) | 204,522 | (3,2) | 150,850 |
| FG-T | (1,2) | 79,908 | (3,2) | 136,644 | (3,3) | 205,756 | (3,2) | 151,914 |
| FG-G | (1,2) | 79,943 | (3,2) | 135,513 | (3,3) | 204,168 | (3,2) | 150,297 |
| FGs-G | (1,3) | 80,199 | (3,2) | 136,013 | (3,3) | 204,849 | (3,2) | 150,676 |
| Cent-G | (1,2) | 79,805 | (3,2) | 135,926 | (3,3) | 204,512 | (3,2) | 150,583 |
| FG-C | (1,2) | 80,172 | (3,2) | 135,793 | (3,3) | 204,944 | (2,2) | 150,839 |
| FG-R | (1,2) | 80,047 | (3,2) | 135,378 | (3,3) | 203,946 | (3,2) | 150,200 |

In addition to information criteria, Table 2 summarises the estimated decay parameters along with their 95% confidence intervals. Across regions, the estimates for each distance measure were comparable when not using the centroid approach. The similarity in decay parameter estimates and the selected seasonality orders indicate that an application of those models to all 401 spatial units jointly is an interesting alternative. For the two distance measures based on the geodesic distance between individuals — the beeline and gravity model — the decay estimates are between 1.4 and 1.6, which aligns with the findings of Brockmann et al. [2006].

**Table 2.**
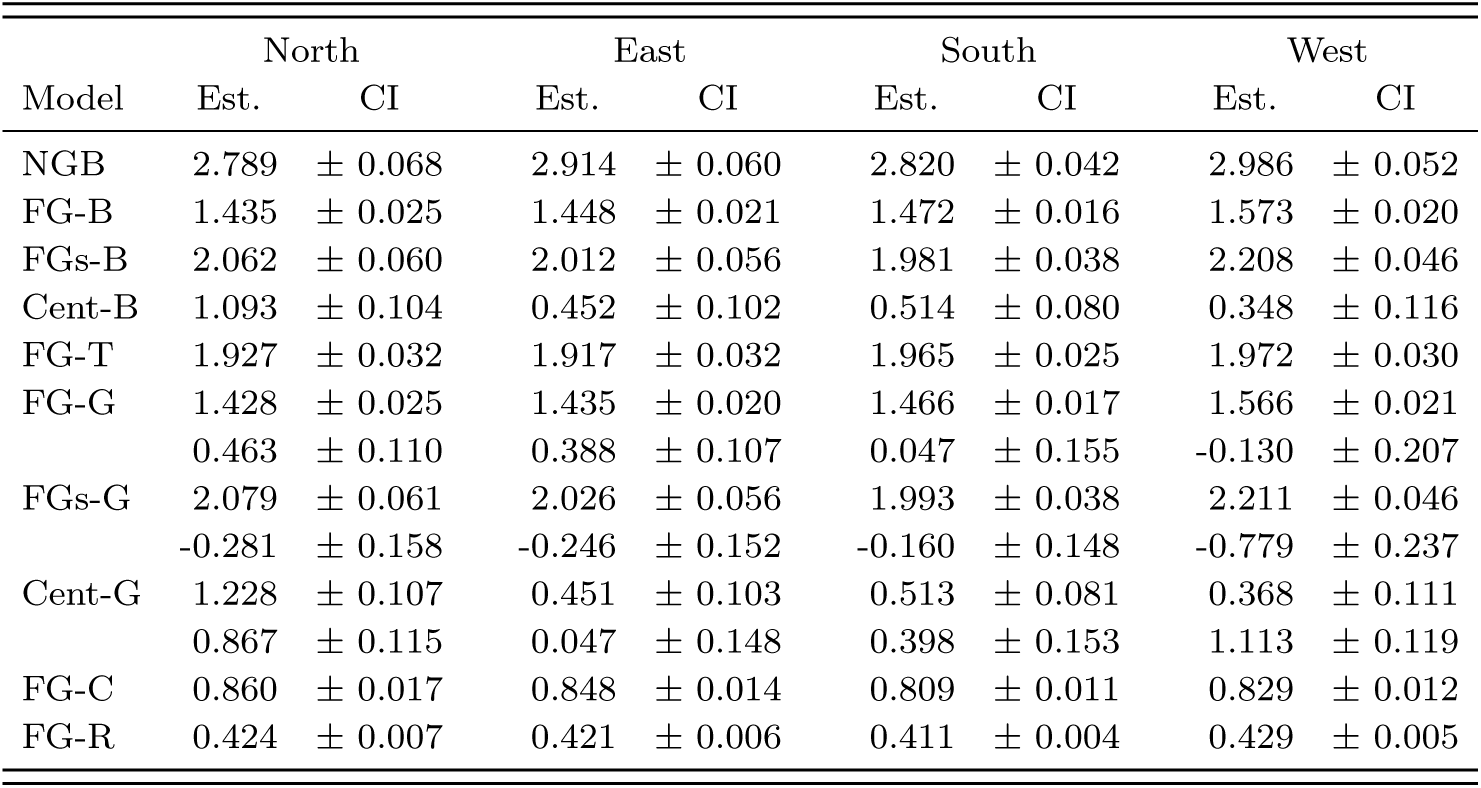
Estimated decay parameters and 95% confidence intervals from fitting to complete data.

For the gravity models, we saw that some estimates for the second decay parameter were negative, implying that a transmission from an infected person living in a less densely populated districts was less likely to a district with high population density. This counterintuitive result emphasises a limitation of the gravity model and other models with asymmetric weights. The transmission risks are based on the distance between an infected person and a destination the person might travel to. However, transmission can also occur when a susceptible person travels to a destination of an infected person. Accounting for those transmission paths is an interesting and necessary future extension.

### 3.2. Forecast Performance

We compared forecast performance across methods, regions, and three different scores: the Weighted Interval Score (WIS) applied to the original forecasts, the WIS applied to log-transformed forecasts and data (lWIS), and the Ranked Probability Score (RPS). The forecast horizons varied between one and eight weeks.

Table 3 presents the aggregated 1-week ahead performance measured with three scores compared to the reference model, with the model exhibiting the lowest (i.e. best) relative score highlighted in bold. The results show that all models performed better in one-week-ahead forecasts than the reference model with no spatial correlation. The centroid gravity approach outperformed fine-grid model approaches in the west regardless of the scoring rule. In the south and east, the circle population approach performed best when a scoring rule on the original scale was used. In the northern region, the centroid gravity approach performed best for scoring rules on the original scale and the travel time approach when applying the WIS on the log scale.

**Table 3.** One week ahead forecast performance - Relative scores compared to the reference model with no spatial autocorrelation.

| Model | North |  |  | East |  |  | South |  |  | West |  |  |
| --- | --- | --- | --- | --- | --- | --- | --- | --- | --- | --- | --- | --- |
|  | WIS | IWIS | RPS | WIS | IWIS | RPS | WIS | IWIS | RPS | WIS | IWIS | RPS |
| NGB | 0.919 | 0.845 | 0.922 | 0.939 | 0.868 | 0.933 | 0.898 | 0.831 | 0.918 | 0.948 | 0.829 | 0.946 |
| FG-B | 0.913 | 0.832 | 0.919 | 0.931 | 0.859 | 0.929 | 0.894 | 0.829 | 0.917 | 0.958 | 0.829 | 0.951 |
| FGs-B | 0.922 | 0.846 | 0.922 | 0.941 | 0.866 | 0.940 | 0.899 | 0.837 | 0.923 | 0.960 | 0.834 | 0.955 |
| Cent-B | 0.909 | 0.835 | 0.920 | 0.935 | 0.878 | 0.936 | 0.900 | 0.825 | 0.916 | 0.981 | 0.842 | 0.971 |
| FG-T | 0.912 | **0.830** | 0.919 | 0.951 | 0.888 | 0.954 | 0.911 | 0.837 | 0.926 | 0.974 | 0.857 | 0.970 |
| FG-G | 0.908 | 0.832 | 0.910 | 0.931 | **0.858** | 0.930 | 0.895 | 0.829 | 0.917 | 0.957 | 0.828 | 0.950 |
| FGs-G | 0.922 | 0.846 | 0.921 | 0.937 | 0.866 | 0.937 | 0.898 | 0.841 | 0.921 | 0.956 | 0.832 | 0.950 |
| Cent-G | **0.897** | 0.833 | **0.903** | 0.934 | 0.877 | 0.935 | 0.894 | **0.822** | 0.910 | **0.921** | **0.818** | **0.921** |
| FG-C | 0.913 | 0.848 | 0.915 | **0.929** | 0.867 | **0.926** | **0.882** | 0.831 | **0.902** | 0.939 | 0.894 | 0.940 |
| FG-R | 0.905 | 0.832 | 0.916 | 0.930 | 0.859 | 0.927 | 0.894 | 0.828 | 0.916 | 0.956 | 0.826 | 0.951 |

Table 4 presents the weighted interval score on a log scale for forecast horizons of 1, 4, and 8 weeks. The methods are ranked according to their performance. In the first block — for the one-week-ahead forecast — we observe the same trends as previously. For the four-week-ahead forecast, the best-performing models were either the circle population measure or the centroid gravity approach. The former was consistently in the best two performing models whereas the centroid gravity model was ranked last for the eastern region. Except for the northern region, most models with spatial correlation did not outperform the reference model without spatial correlation. Only the circle population measure performed better in all four regions.

**Table 4.** Ranking for forecast horizons.

| Rank | North |  | East |  | South |  | West |  |
| --- | --- | --- | --- | --- | --- | --- | --- | --- |
|  | Model | IWIS | Model | IWIS | Model | IWIS | Model | IWIS |
| h = 1 |  |  |  |  |  |  |  |  |
| 1 | FG-T | 12.06 | FG-G | 14.19 | Cent-G | 13.01 | Cent-G | 13.34 |
| 2 | FG-G | 12.09 | FG-B | 14.19 | Cent-B | 13.04 | FG-R | 13.47 |
| 3 | FG-R | 12.09 | FG-R | 14.20 | FG-R | 13.09 | FG-G | 13.51 |
| 4 | FG-B | 12.09 | FGs-G | 14.30 | FG-B | 13.11 | FG-B | 13.53 |
| 5 | Cent-G | 12.10 | FGs-B | 14.32 | FG-G | 13.12 | NGB | 13.53 |
| 6 | Cent-B | 12.13 | FG-C | 14.33 | FG-C | 13.14 | FGs-G | 13.58 |
| 7 | NGB | 12.28 | NGB | 14.34 | NGB | 13.15 | FGs-B | 13.61 |
| 8 | FGs-G | 12.29 | Cent-G | 14.50 | FG-T | 13.23 | Cent-B | 13.73 |
| 9 | FGs-B | 12.29 | Cent-B | 14.50 | FGs-B | 13.23 | FG-T | 13.98 |
| 10 | FG-C | 12.32 | FG-T | 14.67 | FGs-G | 13.29 | FG-C | 14.59 |
| 11 | None | 14.53 | None | 16.53 | None | 15.81 | None | 16.32 |
| h = 4 |  |  |  |  |  |  |  |  |
| 1 | Cent-G | 26.71 | FG-C | 32.77 | FG-C | 31.97 | Cent-G | 34.58 |
| 2 | FG-C | 27.48 | None | 33.61 | Cent-G | 32.97 | FG-C | 36.94 |
| 3 | FG-R | 28.18 | FGs-G | 33.95 | None | 33.39 | None | 37.37 |
| 4 | FG-G | 28.64 | NGB | 33.98 | Cent-B | 33.41 | FG-B | 38.30 |
| 5 | FG-B | 28.79 | FG-R | 33.99 | NGB | 33.50 | NGB | 38.31 |
| 6 | FG-T | 28.82 | FGs-B | 34.02 | FG-R | 33.51 | FG-R | 38.41 |
| 7 | FGs-G | 28.87 | FG-B | 34.02 | FG-G | 33.66 | Cent-B | 38.47 |
| 8 | FGs-B | 29.08 | FG-G | 34.06 | FG-B | 33.68 | FGs-G | 38.59 |
| 9 | NGB | 29.43 | FG-T | 34.63 | FG-T | 33.78 | FG-G | 38.74 |
| 10 | Cent-B | 29.69 | Cent-B | 34.87 | FGs-G | 33.87 | FGs-B | 38.79 |
| 11 | None | 32.25 | Cent-G | 34.97 | FGs-B | 33.90 | FG-T | 39.02 |
| h = 8 |  |  |  |  |  |  |  |  |
| 1 | Cent-G | 37.51 | None | 50.20 | None | 48.09 | None | 51.32 |
| 2 | FG-C | 39.45 | FG-C | 51.33 | FG-C | 50.26 | Cent-G | 53.81 |
| 3 | None | 41.11 | NGB | 52.53 | Cent-G | 53.18 | FG-C | 54.89 |
| 4 | FG-G | 42.32 | FGs-B | 52.75 | NGB | 53.82 | FG-B | 56.48 |
| 5 | FG-R | 43.07 | FG-B | 52.90 | FG-R | 53.87 | FGs-G | 57.81 |
| 6 | FGs-G | 43.50 | FG-G | 53.02 | FG-G | 54.20 | NGB | 57.97 |
| 7 | FG-T | 43.51 | FGs-G | 53.04 | FGs-B | 54.25 | FGs-B | 58.00 |
| 8 | FG-B | 43.87 | FG-R | 53.96 | FGs-G | 54.26 | FG-R | 58.13 |
| 9 | FGs-B | 43.89 | FG-T | 54.36 | FG-B | 54.31 | FG-G | 58.14 |
| 10 | NGB | 44.17 | Cent-B | 55.05 | Cent-B | 54.39 | Cent-B | 58.17 |
| 11 | Cent-B | 45.27 | Cent-G | 55.21 | FG-T | 54.47 | FG-T | 58.89 |

For eight-week-ahead forecasts, we see that the best performing model in three regions is the approach without spatial correlation. The circle population model was in the top three performing models in all four regions. The poor performance of spatial models could be due to the complex nature of epidemic behaviour, where predicting infection counts two months in advance proves to be highly challenging. For longer forecast horizons, it seems that forecasting a typical infection count for the region is the most reliable approach.

### 3.3. Risk Mapping

As an example risk map, we assess the risk for Southern Germany in week 9 of 2019, based on a fitted model that used the beeline distance between individuals as an easily computed measure. This week was the first after the peak of 2019, the most recent year with complete observations. Figure 2 presents the overall risk for grid cells, revealing that areas around Munich exhibited the highest risk due to both the large number of infections in the district and its high population density.

**Fig. 2:**
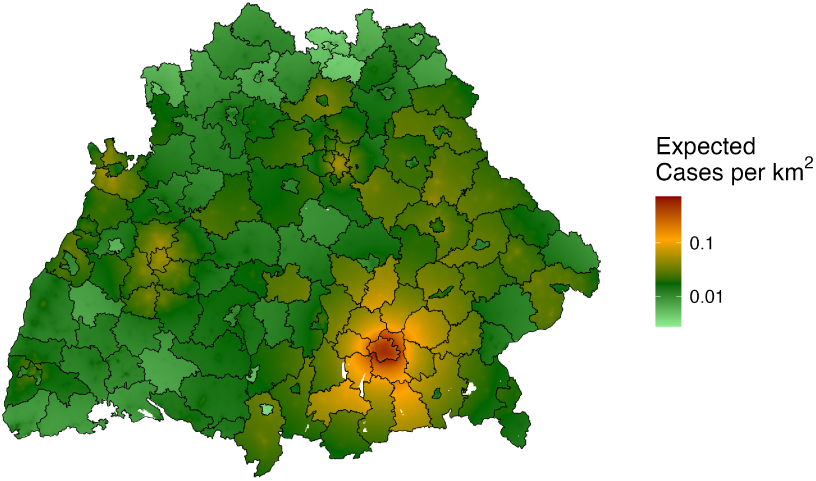
Number of expected cases in Southern Germany in week 9 of 2019 per square kilometer based on beeline distance approach

Figure 3 illustrates the individual risk, showing variation across grid cells within single districts. This detailed risk mapping approach cannot currently be assessed without information on the location of infected individuals within districts. However, evaluating its performance using synthetic data in a simulation study is an interesting future research direction.

**Fig. 3:**
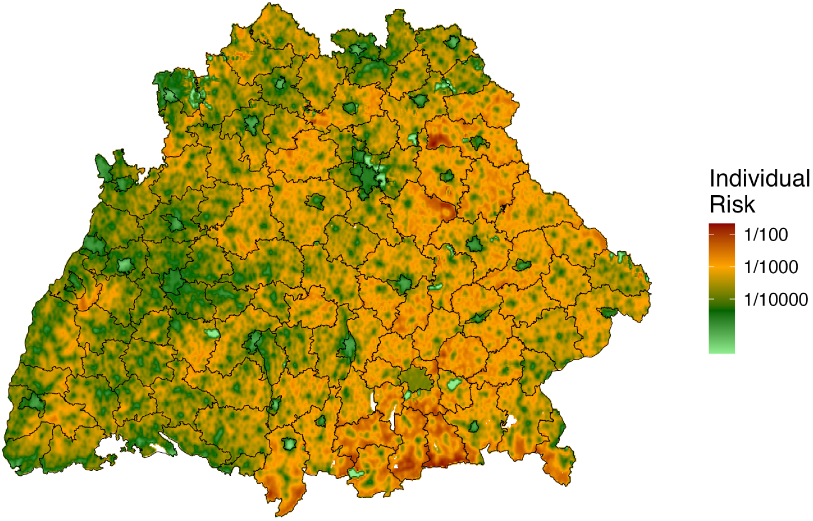
Individual risk of infection in Southern Germany in week 9 of 2019 based on the beeline distance approach

Detailed risk mapping provides a valuable tool for public health planning and intervention. Unlike district-level predictions, which are constrained by administrative boundaries, forecasts on grid cells can be adapted to the specific applications. For example, this approach enables the generation of forecasts for hospital catchment areas, which often do not align with district boundaries. Such tailored forecasts could then help hospital administrators allocating resources more effectively [Meakin and Funk, 2024].

## 4. Discussion

This study assessed the performance of novel spatial approaches for forecasting infection dynamics. By incorporating individual-level distance measures and aggregating to district-level risks, these methods showed robust predictive capabilities. These models often outperformed the model based on neighbourhood orders but they did not consistently outperform the centroid gravity approach in short-term forecasting. Especially the circle population distance measure appeared to have promising forecast performance. Extending this general idea of intervening opportunities not only to include the population structure but other driving factors of human migration is an interesting future research direction.

In addition to their forecasting strength, detailed risk mapping offers a significant enhancement, allowing for a nuanced understanding of spatial heterogeneity and highlighting areas at higher risk both in absolute terms and on a per-capita basis. These insights extend the usefulness of the models beyond administrative boundaries.

Despite these advances, this article also highlights that there is still much to explore. The application considered only one disease and one country. Assessing the robustness of the new method when applied to other diseases or countries can and should be addressed in future research.

The proposed methods are not without their challenges. The estimation procedures are sensitive to the choice of starting values, which can affect convergence and stability. Additionally, the preparation required for estimation is computationally intensive, particularly when dealing with large datasets or high-resolution spatial grids. While the new approaches generally performed well, their forecast performance was comparable to the established neighbourhood order-based method in certain scenarios.

There are numerous directions for future research to address these limitations and expand the utility of the proposed methods: Incorporating alternative distance measures into radiation models could provide insights into how spatial interactions vary under different assumptions. The validity of current assumptions should be investigated in more detail, such as the choice of the Lognormal distribution or translating distances to contact intensity with a power law. More flexible approaches for the latter, such as splines, could be an interesting alternative.

Similar to the detailed risk map, it would be an advantage for retrospective analysis and forecasting to identify clusters or hotspots invariant to district boundaries. Generally, the detachment from the district boundaries in modelling provides more flexibility for applications where district boundaries changed during the observation period. Whether the new approach is robust to such boundary changes can be assessed in a simulation study.

Incorporating additional covariates could offer a more comprehensive understanding of transmission dynamics and risk factors. For example, the age and gender structure of districts could be incorporated in the log-linear predictors or the definition of the weight matrix.

By addressing these areas, future work can build on the foundation laid in this study to develop even more accurate, flexible, and interpretable models for forecasting and risk assessment in epidemic contexts.

### A. Motivating Examples

Using the order of neighbourhood as a measure of closeness between district is simple, but entails pitfalls practitioners need to be aware of. To highlight some, consider the examples shown in Figure 4.

**Fig. 4:**
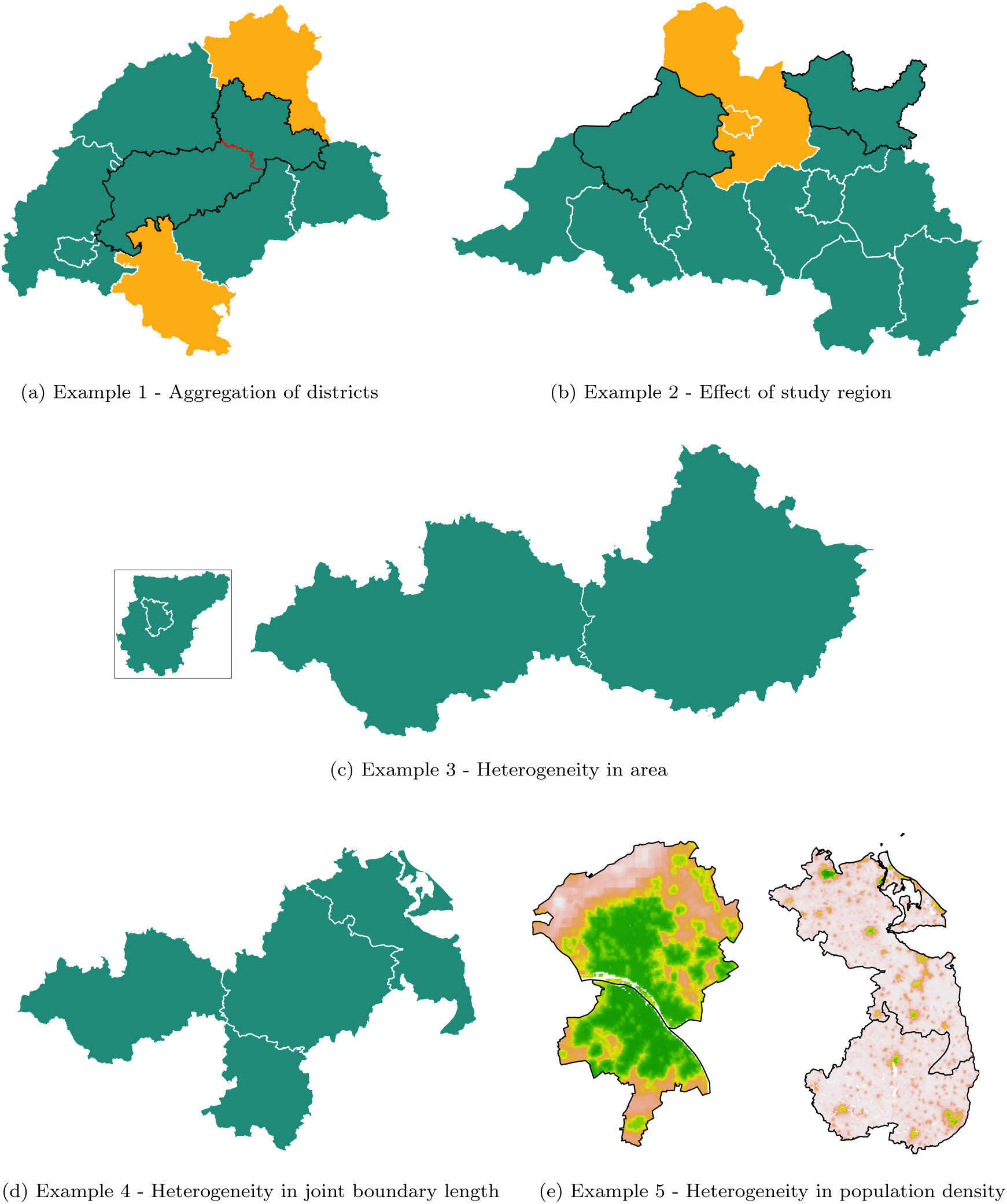
Comparison of different heterogeneity effects

**Fig. 5:**
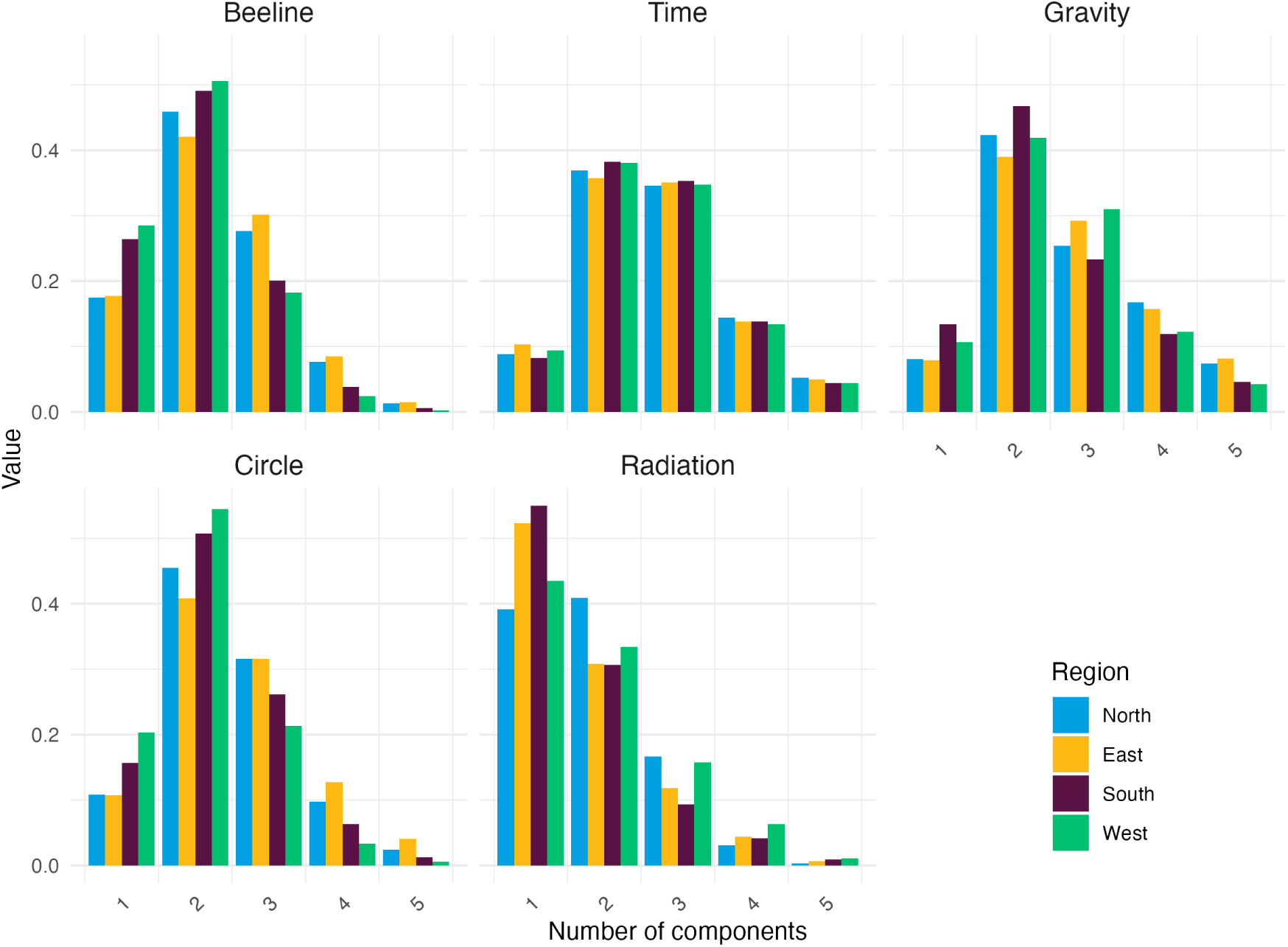
Number of mixture components split by distance measure and region

For some applications, there are reforms in the district boundaries during the observation period. In our application, the districts Göttingen and Osterode am Harz were aggregated in 2016. Figure 4a shows in black outline the current district and in red the former boundary. For the two spatial units highlighted in yellow, the order of neighbourhood changed due to the aggregation and thus how we quantify the transmission risk between the two districts. For us, district reforms have not been problematic, as the number of new infections is reported on the basis of the current districts. However, that is not always the case for other applications. Simple district aggregations can be accounted for by summing up the number of cases in retrospect, but it is not straightforward how to deal with a district being split when only the total number of cases is available for times before the split.

Another example for the sensitivity of neighbourhood orders as distance measure is how we decide for an area of application. Figure 4b shows districts in West Germany (green) and districts in Nothern Germany (blue). In the application we analyse the data separately. For the pair of spatial units highlighted in black outline, Steinfurt and Minden-Lübbecke, the order or neighbourhood is four. If we extend to study area and also include districts in Northern Germany, the order is only two, while most other orders remain the same. Thus, the transmission risk between the two districts is evaluated differently compared to other pairs of districts.

Figure 4c shows two pairs of two districts each both with a neighbourhood order of one. Hence, we would assume similar transmission risk between them. However, the pair on the right is larger in area compared to the pair on the left, where one districts is fully surrounded by the other. It is plausible that the transmission risk between the left pair is more likely than for the right pair. While on the left, individuals live at most 32 kilometers apart with an average of around 13, two individuals from the right pair may live up to 198 kilometers apart with an average of 106.

Example 4, shown in Figure 4d, displays four districts in Eastern Germany. The two districts at the bottom left share only a short border, while the other two districts’ border is substantially longer. Transmission risk is again assumed similar when using neighbourhood orders.

Finally, example 5 in Figure 4e shows a comparison of two pairs where the colours give the population density on a log scale. On the left, we see Mainz and Wiesbaden, to urban districts where the population of both is clustered near their border. On the left, we see the rural districts Uckermark and Vorpommern-Greifswald. The population is more spread out with the most densely populated are in the more northern district is location far away from the border. Such differences are not accounted for when basing transmission risk on the order of neighbourhood.

### B. Weights and Derivatives

#### B.1. Derivation of Weights

In the following, the derivatives of weight matrices are summarised. Given a distance measure *D_ij_*, we define unnormalised weights as the expectation 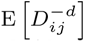 regardless of what *D_ij_* measures, the beeline distance, route time or radiation measure between individuals. If the power law shall be truncated, raw weights are the expectation 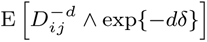, where ”∧” is the minimum operator. The distribution of *D_ij_* is assumed to be either a Lognormal distribution, a truncated Lognormal distribution or a mixture of multiple (truncated) Lognormal distributions. Since truncation boundaries account for the minimum and maximum distance, they are assumed to be the same for all mixing components, if considered. All distributions considered are special cases of a mixture of truncated Lognormal distributions. Further, the regular power law is a limiting case of the truncated power law for *δ* → 0. Thus, we summarise derivatives of raw weights for the most general case, however, we ignore mixture distributions to simplify notation.

Let *W* be a weight such that

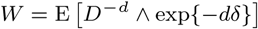

where *D* is the distance measure, *d*and *δ* are parameters. *D* follows a truncated Lognormal distribution with parameters *µ*, *σ*^2^ that takes values between exp(*l*) and exp(*u*). If *l < δ < u*, we split above expectation

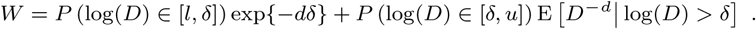

Then, the probabilities are

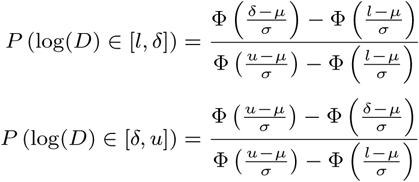

To find the conditional expectation, we see that *D^−d^*| log(*D*) *> δ* follows a truncated LogNormal distribution with parameters −*dµ* and *d*^2^*σ*^2^ truncated between −*du* and −*dδ*. Thus, according to Wang et al. [2012], the expectation is

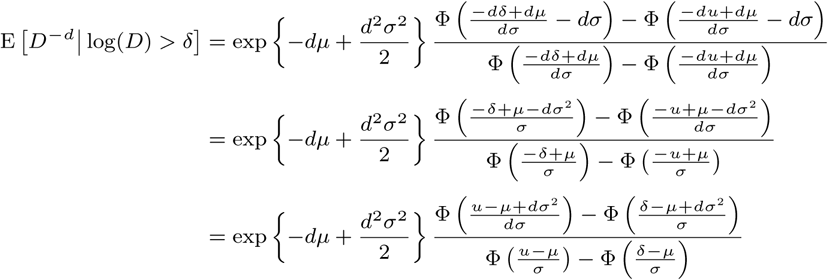

To sum up, we have

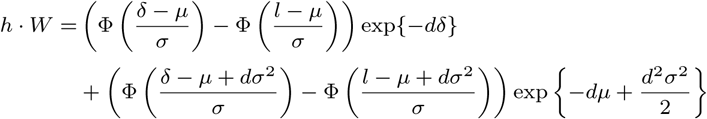

where 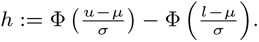

So far, we have restricted to the case *l* ≤ *δ* ≤ *u*. Above findings can be generalised by defining

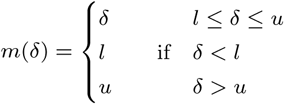

Finally we have weights for the general case in dependence of parameters *δ*, and *d*

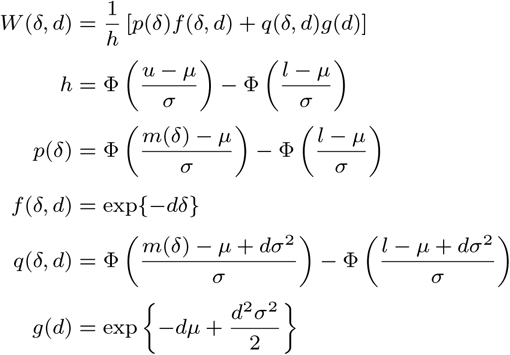

#### B.2. Derivatives of Weights

Above weights are implemented to be used in the *surveillance* package. For numerical optimisation, it is beneficial to use its first two derivatives. To simplify notation, let *f* ^(*d*)^(*δ, d*) and *f* ^(*dd*)^(*δ, d*) be the first and second partial derivatives of *f* (*δ, d*) with respect to *d* and likewise for functions *p*(*δ*)*, q*(*δ, d*) and *g*(*d*) as well as for derivatives with respect to *δ*. Then, we have outer derivatives of *W* (*δ, d*)

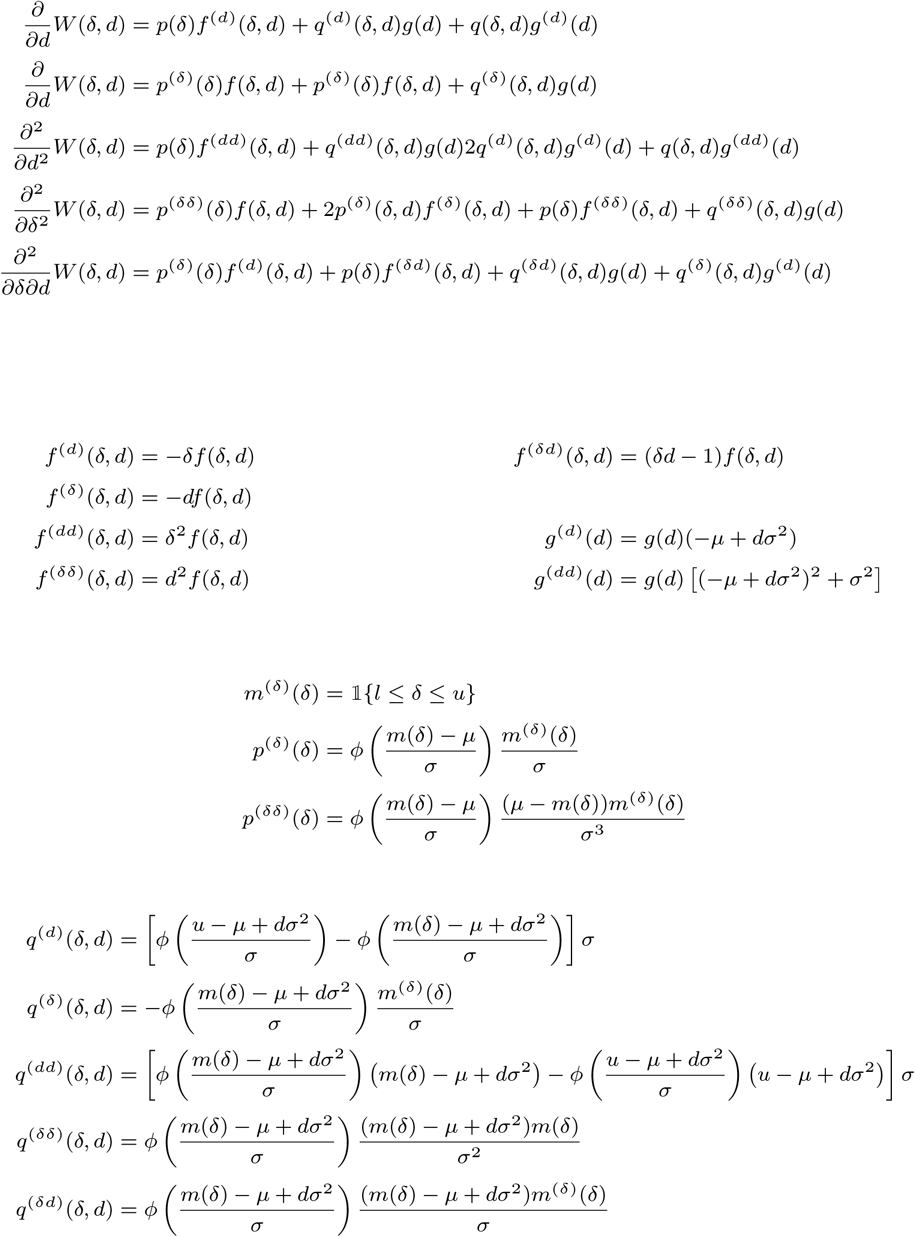

#### B.3. Gravity Model Extension

In case we include gravity effects in the weights, we use that (*D, P* )*^′^* is assumed to follow a mixture of bivariate Lognormal distributions where *D* is a distance measure and *P* is the ratio between population at the origin and destination. Again, for the sake of simplicity, weights are derived ignoring a potential mixture distribution. In that case, we have

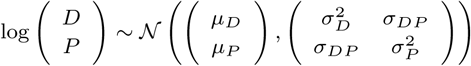

Raw weights are defined as *G* := E *D^−d^*^1^ *P ^d^*^2^. By assumptions made, *D^−d^*^1^ *P ^d^*^2^ follows a Lognormal distribution with expectation

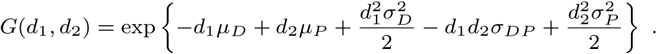

It derivatives with respect to *d*_1_ and *d*_2_ are summarised as

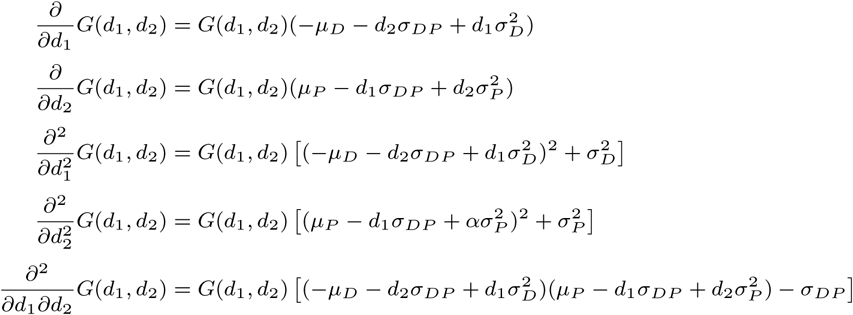

#### B.4. Weight Transformations

From raw weights, whether they are from a distance measure or including gravity effects, we consider three different transformations of weights: 1) normalising the rows of the weight matrix 2) using the parameters on a log scale and 3) scaling the columns. When applying these transformations, the first and second derivatives need to be transformed as well. In the following let *W* be the matrix of weights before applying a transformation and *W* ^(*d*)^ as well as *W* ^(*dd*)^ its first and second derivatives with respect to *d* respectively. Likewise derivatives are defined for other parameters *δ* or *d*_2_ and cross-derivatives.

##### Row normalisation

When the matrix of raw weights is row normalised, we first compute the row sums as *S* = *W* 1*_n×n_*, where 1*_n×n_* is an *n* × *n* matrix of ones. This results in a matrix that has the same dimension as *W* and contains the corresponding row sums. Thus, the normalised weight matrix can be expressed as *W* ⊘ *S*, where ⊘ is the Hadamard division operator. Its derivative with respect to *d* is simply

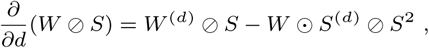

where ⊙ is the Hadamard product. Derivatives with respect to *δ* or *d*_2_ are found analogously. The second derivative is

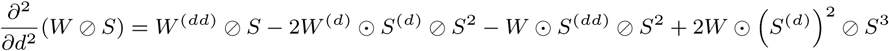

The cross derivatives with respect to *d*_1_ and *d*_2_ are

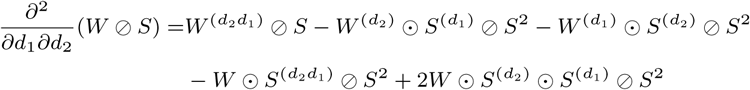

##### Log Transformation

If weight matrices are computed using parameters on a log scale, i.e. parameters is transformed by the exponential function before inserted into the weight function, derivatives are simpl

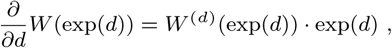

second derivatives

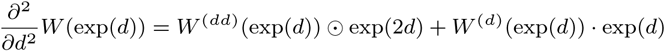

and cross-derivatives

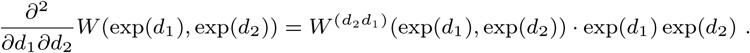

##### Scaling

If a weight matrix is scaled column-wise, i.e. *W_ij_* 1→ *W_i,j_* · *c_j_* for constants *c_j_*, we can simply define *C* as the diagonal matrix containing *c_j_*. Then, *W* is transformed to *W* · *C* and likewise all derivatives are simply multiplied by *C*.

## Data Availability

All data used are publicly available and summarised at https://zenodo.org/records/15600537. General functions to apply the methods are available in a toolbox at https://github.com/ManuelStapper/FGSIM. Application-specific code is available at https://github.com/ManuelStapper/FGSIM_Application.

https://survstat.rki.de/

https://dx.doi.org/10.5258/SOTON/WP00645

## 6. Author contributions statement

M.S. conceived the study, developed the methodology, implemented the software, conducted analyses, and wrote the initial draft. S.F. provided supervision, contributed to study design refinement, suggested model variants, and revised the manuscript. Both authors reviewed and approved the final version.

## 7. Acknowledgments

This work was supported by funds from Wellcome (210758/Z/18/Z) and the National Institute for Health and Care Research (NIHR) Health Protection Research Unit (HPRU) in Health Analytics and Modelling. The authors acknowledge the use of Claude Sonnet 4 (Anthropic) for formatting and proof reading. All AI-generated content was thoroughly reviewed, fact-checked, and revised by the authors. The final analysis, conclusions, and academic interpretations are entirely the authors’ own work.

